# Application of AI generated text-to-video in medical education: Systematic review

**DOI:** 10.1101/2025.02.03.25321572

**Authors:** Yaara Artsi, Vera Sorin, Benjamin S. Glicksberg, Panagiotis Korfiatis, Girish N Nadkarni, Eyal Klang

## Abstract

**Background:** Traditional medical education often struggles to simplify complex concepts for both healthcare professionals and patients. AI-generated text-to-video technologies are emerging as tools to enhance medical education by transforming intricate medical content into accessible visual formats. This systematic review aims to evaluate the current literature on the application of AI-generated text-to-video technologies in medical education.

**Methods:** A comprehensive search was conducted in MEDLINE/PubMed, Google Scholar, Scopus, Cochrane Review, and Web of Science for studies published up to January 2025. The search targeted AI-generated text-to-video applications in medical education and patient engagement. Studies were screened based on predefined inclusion and exclusion criteria, and data were extracted independently. The risk of bias was assessed using the QUADAS-2 tool, and the review adhered to PRISMA guidelines.

**Results:** Out of 103 identified studies, 5 met the inclusion criteria. Four studies focused on patient education, and one on physician training. Applications spanned various specialties, including ophthalmology, neurosurgery, plastic surgery, and stroke rehabilitation. AI-generated videos showed potential to improve patient understanding, engagement, and confidence. However, limitations included data biases, content inaccuracies, lack of comparison with traditional methods, and variability in user technological proficiency.

**Conclusion:** AI-generated text-to-video technology holds promise for advancing medical education by improving engagement, enhancing learning outcomes, and facilitating patient understanding. Nevertheless, challenges related to data accuracy, algorithmic bias, ethical concerns, and equitable access must be addressed. Ongoing research, validation studies, and ethical oversight are essential to ensure the safe, effective, and inclusive integration of this technology in medical education.

## Introduction

Mastering medicine requires a unique combination of scientific knowledge, emotional intelligence, compassion, curiosity, creative and critical thinking skills [1,2]. As a result, the process of becoming a physician is long and arduous, and the medical education field is continuously developing [3,4]. Naturally, teaching and understanding medical decision-making and procedures is intricate [5].

According to the patient-centered care approach, patients should be active participants in their healthcare [6, 7]. However, medical terminology and reasoning often remain obscure to the average patient [8, 9]. Using complex medical language can lead to confusion, anxiety and additional stress for patients [10]. This also increases the burden on physicians who need to bridge these gaps in communication.

Artificial intelligence (AI) and natural language processing (NLP) technologies continue to evolve [11, 12]. The latest ground-breaking development is AI generated text-to-video technology such as openAI’s Sora and Synthesia [13, 14]. These tools create videos automatically by learning from vast data sources. This technology can be utilized to enhance medical education for both doctors and patients, by transforming complex medical concepts into accessible visual information [15. 16]. However, the implementation of this new technology might pose several risks and challenges such as data bias, inaccuracies, misuse and equitable access within the global healthcare community [17, 18]. These factors must be addressed to ensure safe, effective, and ethical application.

The aim of this study is to systematically review the current literature on applications of AI generated text-to-video for medical education.

## Methods

### Literature search

We conducted a search to identify studies describing application of AI generated text-to-video for medical education. We searched MEDLINE\PubMed, Google scholar, scopus, cochrane review and web of science for papers published up to January 2025. The search was conducted using the following Boolean operators:

(“text-to-video generation” OR “AI video generation”) AND (“patient education” OR “medical education” OR “health education” OR “patient engagement” OR “health communication”) AND (“medicine” OR “healthcare” OR “clinical practice” OR “hospital” OR “public health”)

We checked the references lists of selected publications for more relevant papers. Sections as ‘Similar Articles’ below articles (e.g., PubMed) were also inspected for possible additional articles.

Ethical approval was not required, as this is a systematic review of previously published research, and does not include individual participant information. Our study followed the Preferred Reporting Items for Systematic Reviews and MetaAnalyses (PRISMA) guidelines [19]. The study is registered with PROSPERO (CRD 42025640042)

### Inclusion and exclusion process

Publications resulting from the search were initially assessed by two authors (YA and VS) for relevant titles and abstracts. Next, full-text papers underwent an independent evaluation by two authors (EK and BSG).

### Inclusion Criteria

The inclusion criteria were as follows: (1) Studies involving medical students, practicing healthcare professionals, or patients in a medical education context. (2) Studies evaluating AI-generated text-to-video technologies used in medical education, such as teaching theoretical concepts, practical skills, or patient education. (3) Studies comparing AI-generated text-to-video tools with traditional educational methods or other digital education tools. (4) Original research such as randomized controlled trials (RCTs), quasi-experimental studies, observational studies, mixed-methods studies, and qualitative research. (5) Studies published in English. (6) Studies published from 2010-01/2025, to focus on the recent development of AI-generated video technologies.

### Exclusion Criteria

The exclusion criteria include: (1) Studies focused on non-medical education or general education without specific applications to healthcare or medicine. (2) Studies that do not involve AI-generated text-to-video technologies, (3) Non-original studies such as opinion pieces, editorials, or commentaries without primary data. (4) Studies published in languages other than English. (5) Abstract-only publications, dissertations, or unpublished theses without access to full data.

Any study in question was discussed among all authors until reaching a unanimous agreement. Risk of bias and applicability were evaluated using the tailored Quality Assessment of Diagnostic Accuracy Studies tool 2 (QUADAS-2). (**Figure 2**.)

## Results

### Study selection and characteristics

The search yielded 103 studies, out of which 53 were duplicates and 45 were excluded due to publication type and irrelevance to the reviews subject (**Figure 1**.) Out of 5 relevant studies, 4 (80%) detailed utilization of AI generated text-to-video for the purpose of patient education, and one (20%) discussed the technology utilization for physician medical training. Two studies evaluated applications in ophthalmology, one in neurosurgery, one in plastic surgery, and one in neurosurgery. The studies were conducted in various medical fields and applied different AI tools (**Table 1**).

**Figure 1.**
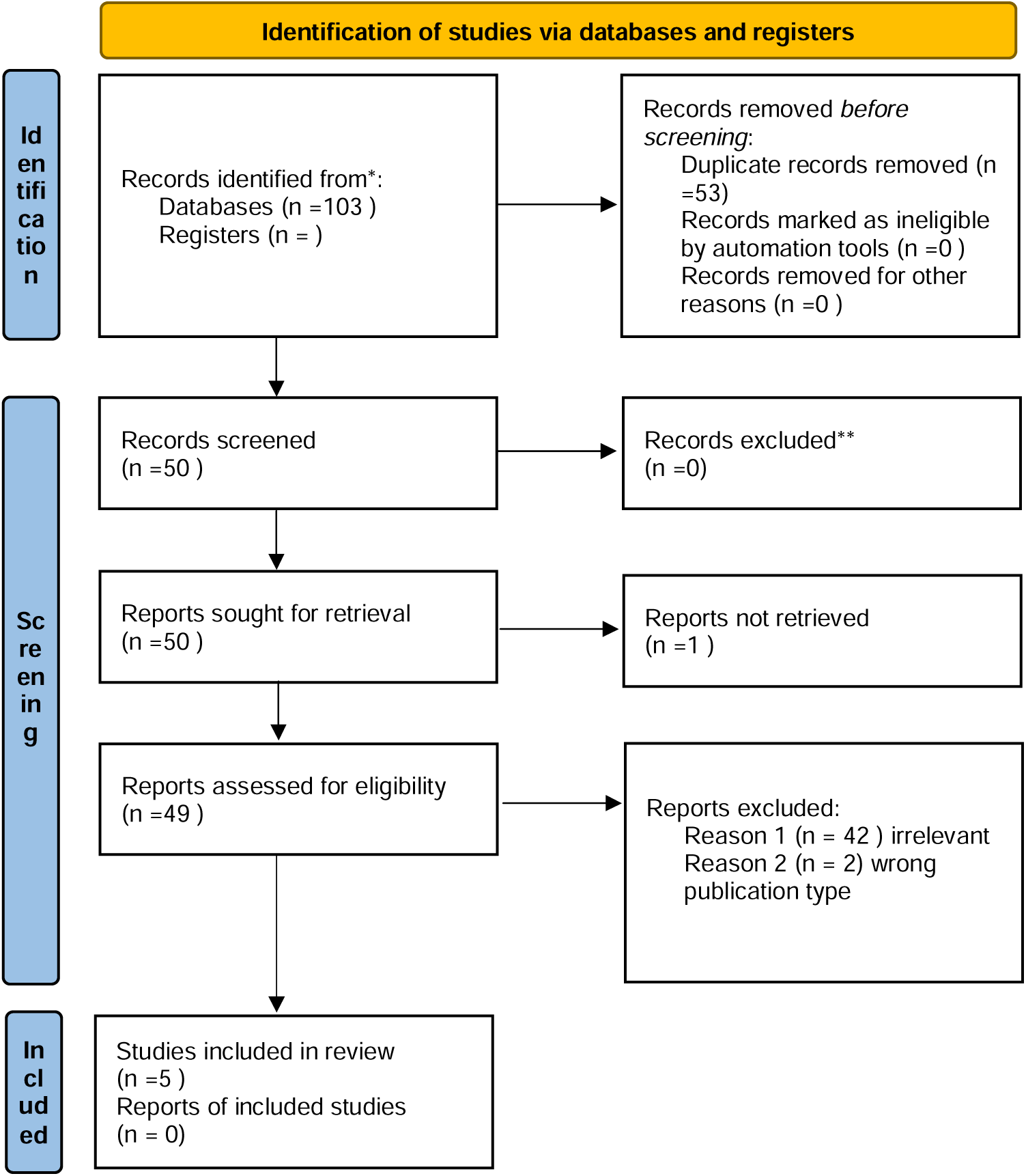
Flow diagram of inclusion and exclusion process

**Figure 2.**
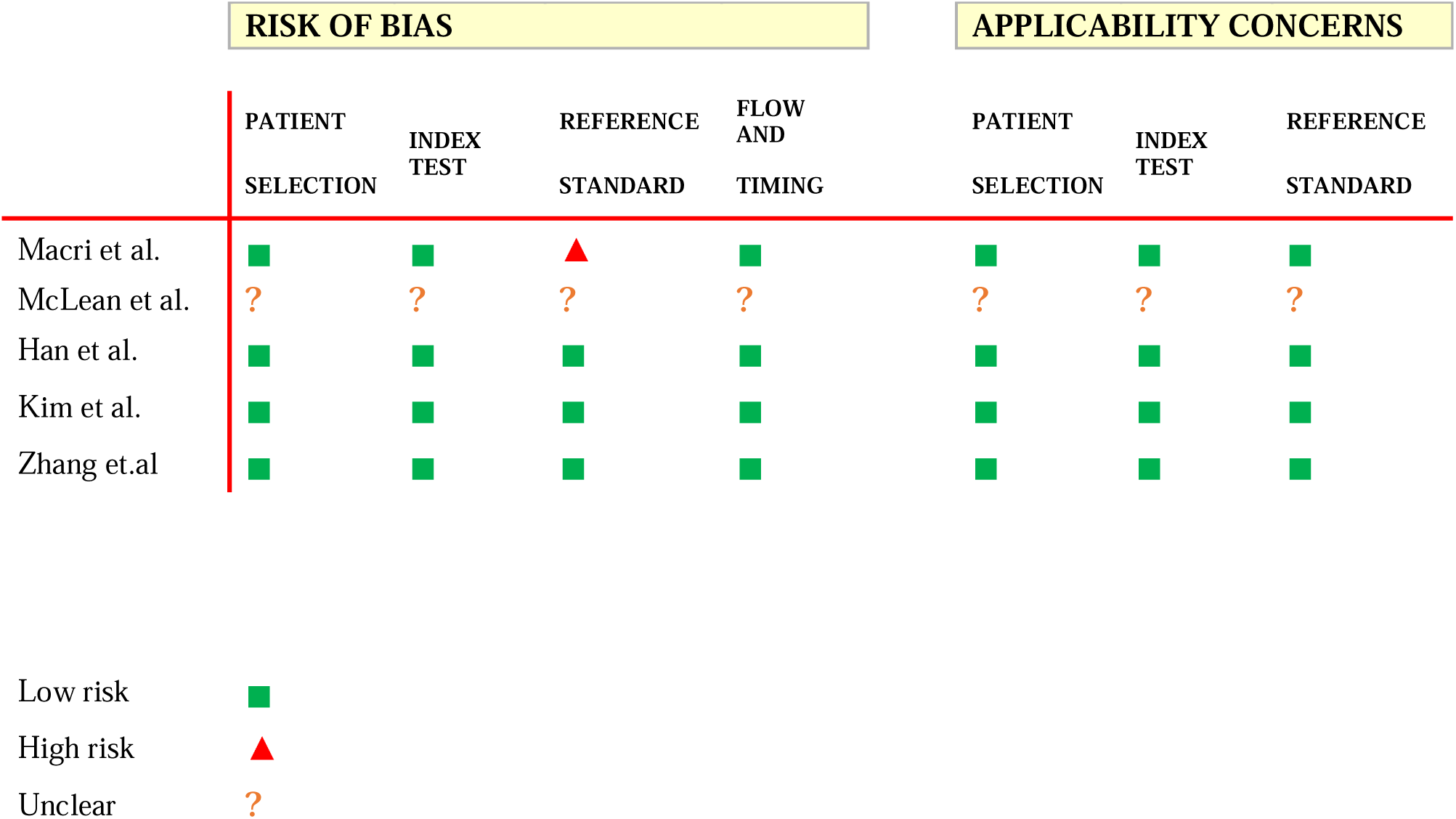
Risk of bias and applicability assesment

**Table 1.** Studies included in the review and their key features.

| Authors | Date of publication | Journal | Intervention | Medical field |
| --- | --- | --- | --- | --- |
| Macri et al. | 09/2024 | Ophthalmic Research | Patient education | Ophthalmology |
| McLean et al. | 09/2024 | International Journal of Computer Assisted Radiology and Surgery | Physician education | Neurosurgery |
| Han et al. | 10/2024 | Cureus | Patient education | Ophthalmology |
| Kim et al. | 10/2024 | PRS Global Open | Patient education | Plastic surgery |
| Zhang et.al | 10/2024 | Clinical Nutrition | Patient education | ENT/Nutrition |

### Descriptive summary of results

*Macri et al.* [20] examined patient response to an AI-generated presenter in educational videos concerning postoperative care in face-down positioning after vitreoretinal surgery. The participants filled a pre-video survey and the 6-item Spielberger State-Trait Anxiety Score (STAI-6). The post-video survey included rating the quality of the video using the Global Quality Score and questions regarding the participants perceptions of the AI video. In addition a free-text response was also collected.

Post-video assessment overall 11 patients rated the quality of the video as “excellent” (73%). 9 patients (60%) understood, felt at ease and trust the presenter. 7 patients (47%) would watch more videos with the presenter, and support using AI generated presenters in the video. In the free-text feedback, patients reported overall positive experience with the AI presenter (**Table 2**.). Despite promising results, there was no comparison with traditional methods of information delivery, such as human presenters or written materials, making it difficult to determine the relative effectiveness of the AI-generated presenter (**Table 3**.).

**Table 2.** Summary of parameters used in the studies.

| Study | AI tool | Control tool | Performance metrics | Prompt | General results |
| --- | --- | --- | --- | --- | --- |
| Macri et al. | Synthesia | N/A | Surveys<br>STAI-6 score | N/A | >50%<br>Agreed the AI video is clear, helpful and reassuring |
| McLean et al. | Sora | Traditional training methods | Technical skill<br>Error rate<br>Procedure completion time<br>Cognitive load assessment<br>Decision making quality<br>Feedback from trainees<br>Peer and mentor reviews<br>Pre- and post-assessment<br>Continuous monitoring | "A video simulation of the step-by-step process of resecting a brain tumor, including the approach, craniotomy, and tumor removal" | Immersive simulations for skill development.<br>Exposure to a diverse range of surgical scenarios.<br>Standardized, objective methods for assessing and providing feedback on trainee competencies. |
| Han et al. | InVideo/ClipTalk/EasyVid | Gold standard control videos from the American Academy of Ophthalmology (AAO) | 3-point scale of four-category quality assessment:<br>Image accuracy<br>Script accuracy<br>Image clarity | "Create a patient-centered educational video depicting a step-by-step guide on what to expect during LASIK, PRK, or SMILE surgery. Be as anatomically and | Medical accuracy:<br>Control > InVideo, EasyVid, and ClipTalk for LASIK, PRK and SMILE (p<0.005)<br>Video quality and resolution:<br>no significant differences were found |
|  |  |  | Script alignment with the video | surgically accurate as possible." | ($p > 0.05$)<br>Script accuracy:<br>Control > ClipTalk for the SMILE instructional video<br>($p < 0.05$)<br>Script alignment:<br>AI-generated videos had comparable alignment between the script and the images being displayed compared to the control videos<br>Total score:<br>control videos outperform all three AI text-to-video generators |
| Kim et al. | Synthesia | Automated text-based chatbot integrated with ChatGPT 3.5 | 1–7 Likert scale rating 16-item survey | N/A | No difference in perceptions between the two platforms<br>participants preferred the VideoBot over the traditional chatbot (63.5% versus 28.1%) |
| Zhang et.al | AI-VG | Usual care treatment:<br>Lip, tongue and chin exercises | swallowing function measured by (GUSS)<br>Standard Swallowing Assessment (SSA)<br>Functional Oral Intake Scale (FOIS)<br>Mini-Nutritional Assessment Short Form (MNA-SF)<br>Swallowing Quality-of-Life Questionnaire (SWAL-QOL)<br>Technology | N/A | AI-VG group had significantly improved GUSS scores ($P < 0.001$)<br>AI-VG group had lower SSA scores with no statistical difference ($P = 0.100$)<br>AI-VG group showed an additional increase in the FOIS scores ($P < 0.001$)<br>AI-VG group had a significantly improved MNA-SF score ($P = 0.034$)<br>AI-VG group showed a more significant decrease in SWAL-QOL |
|  |  |  | Acceptance<br>Model (TAM) |  | *(P* < 0.001)<br>Only AI-VG group<br>completed the<br>acceptance and<br>satisfaction<br>questionnaires<br>Score 73.00 [72.00–<br>74.00] |

**Table 3.** Limitations found in the studies.

| Study | Technical limitation | Evaluation limitation | Performance limitation |
| --- | --- | --- | --- |
| Macri et al. | N/A | No comparison with traditional methods of information delivery | N/A |
| McLean et al. | Implementing transformer architecture requires significant processing power and specialized hardware | No standardized metrics to evaluate the quality and educational value of AI-generated simulations | Generated scenarios may lack the unpredictability and complexity of actual surgical situations |
| Han et al. | N/A | No comparison between AI-generated videos and traditional educational methods | non-surgical ophthalmic video clips<br>random clips from different surgical specialties<br>completely factitious details |
| Kim et al. | No flexibility in asking the VideoBot other questions, due to incomplete integration of ChatGPT into the Synthesia.ai platform | N/A | Difficulty for some patients to process information in a video setting as opposed to reading at their own pace |
| Zhang et.al | N/A | Different educational background can affect comfort and competency with technological interfaces | N/A |

*McLean et al.* [21] aim to assess how transformer-based architectures, specifically OpenAI latest video generation model, Sora, can revolutionize neurosurgical education by providing realistic surgical simulations. These simulations are intended to enhance the learning experience and proficiency of neurosurgical trainees in complex procedures. The study involves compiling a dataset of neurosurgical procedures to train the Sora model, focusing on generating accurate, high-fidelity simulations. The performance metrics focus on skill acquisition, accuracy of procedural execution, and decision-making capabilities. They indicate potential benefits of generative video modeling technologies in neurosurgical training, providing immersive simulations that could aid in skill development and enhance exposure to diverse surgical scenarios. (**Table 2**.) Several limitations were discussed, such as Data Requirements, developing realistic simulations necessitates extensive datasets of neurosurgical procedures, which may be difficult to compile due to privacy concerns and the rarity of certain surgeries. Implementation concerns due to significant processing power and specialized hardware requirements. In addition the need to establish standardized metrics to evaluate the quality and educational value of AI-generated simulations. (**Table 3**.)

*Han et al.* [22] created educational videos for laser-assisted in situ keratomileusis (LASIK), photorefractive keratectomy (PRK), and small incision lenticule extraction (SMILE). They utilized three AI text-to-video platforms; InVideo, ClipTalk and EasyVid. For evaluation of the AI performance, A three-point grading system was used to compare videos in terms of “image accuracy,” “script accuracy,” “image clarity,” and “script alignment.“

For medical accuracy of images used in each instructional video, the control videos outperformed InVideo, EasyVid, and ClipTalk for LASIK (p<0.005), PRK (p<0.005), and SMILE (p<0.005). In terms of script accuracy, the control video had a higher script accuracy score than ClipTalk for the SMILE instructional video (p<0.05). Additionally, each AI platform made errors in their scripts with varying degrees of severity. For script alignment, the AI-generated videos had comparable alignment between the script and the images being displayed compared to the control videos, with the exception of the SMILE video generated by the control, which had better alignment than InVideo (p<0.05). The total score showed the control videos outperformed all three AI text-to-video generators for LASIK, PRK and SMILE (p<0.005) (**Table 2**.).

The study did not include a comparison between AI-generated videos and traditional educational methods, such as in-person consultations or standard video presentations, limiting the ability to evaluate relative effectiveness. Also, some inaccuracies were mentioned, InVideo platform used a combination of non-surgical ophthalmic video clips, including slit lamp examinations and random clips from different surgical specialties. Furthermore, EasyVid and ClipTalk almost always displayed AI-generated images that were related to ophthalmic surgery but included completely factitious details such as a laser emitting directly from an overhead surgical spotlight. (**Table 3**.).

*Kim et al.* [23] evaluated patients’ preferences and perceptions regarding two types of AI virtual assistants, a traditional text-based chatbot and a human-like AI VideoBot generated by synthesia. A total of 396 responses were gathered from women aged 18 to 64 years old. Most of the participants (73%) were aged between 25 and 34 years. The women were provided with interactions from both the traditional chatbot and the AI VideoBot, each delivering identical information about breast reconstruction common questions and procedures. Subsequently, they completed a survey assessing their preferences, perceived effectiveness, engagement levels, and overall satisfaction with each virtual assistant.

They found that when comparing the VideoBot and chatbot, perceptions of truthfulness (*P* = 0.5965), believability (*P* = 0.4834), expertise (*P* = 0.6208), ease of use (*P* = 0.2253), and safety (*P* = 0.2461) were not significantly different. However, the majority of participants preferred the VideoBot over the traditional chatbot (63.5% versus 28.1%), stating that they found the VideoBot to be more captivating than the text-based chatbot (**Table 2**.).

An interesting observation was noted in one participant stating they preferred the traditional chatbot since it was easier to take the time to read and comprehend the given information as opposed to the “rushed” information delivery of a VideoBot. Moreover, the participants were only allowed to ask certain common questions to the VideoBot with no flexibility in asking other questions. This was due to the technological limitations of not being able to fully integrate ChatGPT into the Synthesia.ai platform, which prevented the ability of an open discussion, which was given in the traditional chatbot experience (**Table 3**.).

*Zhang et al.* [24] investigated the efficacy of an AI-based video game system in treating dysphagia among stroke patients. 84 Participants were randomly assigned to either the AI-VG system group or the usual care group. All participants received training for 30 min per session per day, five times per week for 4 weeks. The primary outcome was change in swallowing function from baseline (T0) to post-intervention (T1) and 1 month follow-up (T2). Secondary outcomes included changes in laryngeal function, oral intake function, nutritional status, and swallowing-related quality of life. The adherence, satisfaction, and acceptance of the two groups were evaluated.

Compared with the usual care group, the AI-VG group showed significantly improved swallowing function, with a mean group difference of 4.02 (*P* < 0.001) at T1 and 4.14 (*P* < 0.001) at T2. Oral intake function, nutritional status, and swallowing-related quality of life improved significantly (*P* < 0.001 for overall group × time interaction). Adherence was significantly higher in the intervention group than in the control group (*P* < 0.001). The intervention group had higher levels of acceptance and satisfaction of AI-VG (73.00 [72.00–74.00]). No significant difference was observed in laryngeal function (*P* > 0.05) (**Table 2**.).

A notable limitation mentioned is the lack of baseline in technological proficiency among participants. This difference can affect the degrees of comfort and competency with technological interfaces, influencing engagement and adherence to treatment (**Table 3**.).

These studies underscore a range of benefits, limitations, and contexts in which AI text-to-video could be applied. We discuss these findings in the following section.

## Discussion

In this study we aimed to systematically review the current literature on the application of AI-generated text-to-video technology in medical education and its potential to advance the field. Medical education holds the utmost important task of training competent physicians. It also involves effectively communicating information to patients, supporting their understanding and engagement with their care [25].

Following our review of the five included studies, it is evident that the literature on AI-generated text-to-video in medical education is nascent. Among the five studies we reviewed, four focused on patient education [20, 22, 23, 24], while one examined neurosurgical training [21]. Despite the small corpus, their diverse clinical applications—from vitreoretinal surgery to stroke rehabilitation—demonstrate this technology’s versatility. One study showed patients felt more confident and reassured after watching an AI-generated presenter, though it lacked direct comparisons to standard methods [20]. Another proposed neurosurgical training simulations, but noted the need for extensive datasets and standardized metrics before such tools could be routinely used [21]. Research on refractive surgery videos exposed inaccuracies in AI-generated visuals, revealing the importance of human oversight [22]. Meanwhile, comparing an AI VideoBot to a traditional chatbot highlighted the allure of virtual presenters but also underscored technical limits, such as pre-scripted question formats [23]. Finally, a stroke rehabilitation video game boosted patient adherence, yet differences in baseline technological skills shaped overall engagement [24] (**Figure 3**.).

**Figure 3.**
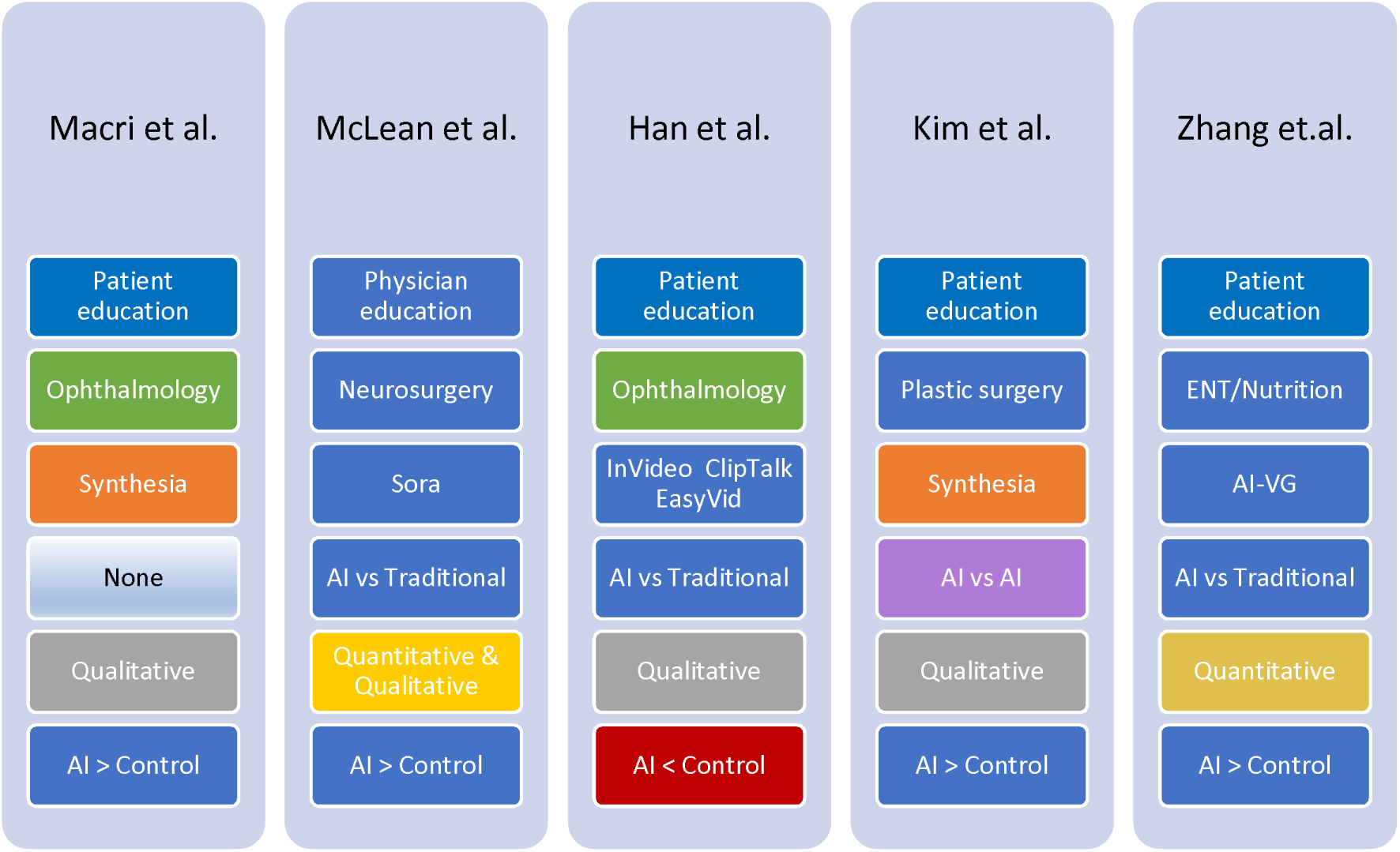
Schematic overview of similarities and differences among the studies

These studies suggest potential gains in engagement, confidence, and learning outcomes. However, they also reveal the need for consistent validation protocols, larger and more diverse study samples, and equitable integration strategies [17, 18, 22]. While promising, AI generated text-to-video requires thorough vetting to ensure accuracy, minimize bias, and expand access across varied clinical settings.

The integration of AI already transformed healthcare, offering innovative solutions for diagnostics, treatment plans, patient management, and education [26, 27]. In medical education, tools like interactive learning platforms, virtual reality simulations, and chatbots have advanced the field [28]. Text-to-video systems could represent the next step in medical education, with the potential to deliver realistic simulations, provide clear visual explanations of medical procedures, and enhance learning experiences and accessibility.

However, integrating AI generated videos into medical education poses challenges and risks. A notable concern is inaccurate presentation of medical information due to errors in the input text or limitations of the AI’s training data [29]. We used the prompt mentioned in Han et al. [22] and applied it to the sora platform. The video generated by sora included spelling errors in the procedure name “PMK” instead of “PRK”, as well as misspelling the word procedure as “proecture” (**Figures 4. and 5.**) In addition AI systems trained on biased data might perpetuate stereotypes or fail to represent diverse patient populations, potentially marginalizing specific groups [30].

Ethical considerations surrounding AI’s role in medical education must also be addressed. AI-generated avatars and videos could be misused to fabricate content, raising concerns about deception and authenticity in medical education [31]. Moreover, sensitive medical information used to generate videos such as patient instructions may be at risk of unauthorized access or adversarial attacks [32, 33].

Another concern is the inability to equally distribute the resources for advanced AI system implementation, especially in low-resource settings. Developing and deploying AI text-to-video systems can be expensive, particularly for institutions with limited budgets; the lack of access to the hardware, internet, or software required to utilize AI-generated video content, can deepen the disparity among different communities [34]. In addition, AI text-to-video systems may not support all languages or dialects, limiting their utility in multicultural or global contexts [35].

It is difficult to replace human interaction and compassion, especially when delivering bad news [36, 37]. We used once more the prompt mentioned in Han et al. [22] and applied it this time to the synthesia platform in order to generate an AI avatar. The result can be shown in the supplementary material (**Supplementary figure 1.**). AI-generated avatars and voiceovers may lack emotional expression or nuance, reducing the effectiveness of videos for sensitive topics like palliative care or mental health. While the studies we reviewed showcase promising results, they also present limitations and challenges that need to be addressed before widespread adoption can occur.

### Mitigation Strategies

To address these risks and challenges several steps can take place, such as accuracy checks, and establishing protocols for validating AI-generated content through professional human review. In order to minimize bias, diverse and representative datasets can be utilized to train the AI models. In terms of data security, implementation of encryption and adherence to privacy regulations can protect sensitive information. For accessibility efforts, future models can be developed with language-inclusive tools for low-resource settings. By proactively addressing these risks and challenges, AI text-to-video technology can be harnessed safely and effectively in medicine and medical education.

### Limitations

Our review has several limitations. First, AI text to video application in medicine and medical education is a new technology, hence, our search did not yield many relevant studies. Future research of the subject will benefit from more studies in this field for better comprehension of AI video integration in medical education.

Second, due to heterogeneity in study design and data, a meta-analysis was not performed. One study, focussed on the theoretical integration of AI text to video application in medical education and suggests framework and synthetic data. Another study compared AI video with AI chatbot, and not with traditional educational methods. One study was at high risk of bias. Additional studies will be needed to further solidify the usefulness of AI videos in medicine, especially in medical education.

## Conclusion

AI-generated text-to-video technology represents an exciting advancement in medical education. It has the potential to enhance learning, improve engagement, and bridge gaps in patient understanding. However, its safe and effective implementation requires attention to challenges such as accuracy, bias and equitable access. Continued research, rigorous evaluation, and ethical oversight are needed to ensure that this technology transforms medical education in a way that benefits both patients and healthcare professionals.

## Data Availability

All data produced in the present work are contained in the manuscript

## Abbreviation

AI: artificial intelligence
NLP: natural language processing
STAI-6: Spielberger State-Trait Anxiety Score 6
LASIK: laser-assisted in situ keratomileusis
PRK: photorefractive keratectomy
SMILE: small incision lenticule extraction
AI-VG: artificial intelligence-based video-game
GUSS: Gugging Swallowing Screen; SSA, Standard Swallowing Assessment
FOIS: Functional Oral Intake Scale
MNA-SF: Mini-Nutritional Assessment Short Form
SWAL-QOL: Swallowing Quality-of-Life Questionnaire
GEE: general estimating equation

## Declarations

### Ethics approval and consent to participate

Not applicable

### Consent for publication

Not applicable

### Availability of data and materials

All data generated or analysed during this study are included in this published article and its supplementary information files.

### Authors’ contributions

YA conducted literature search, data synthesis and wrote the primary manuscript. VS, BSG and PK reviewed and edited the manuscript. GNK and EK contributed in conceptualization, data synthesis and editing of the manuscript. All authors read and approved the final manuscript

## Acknowledgements

Not applicable

---

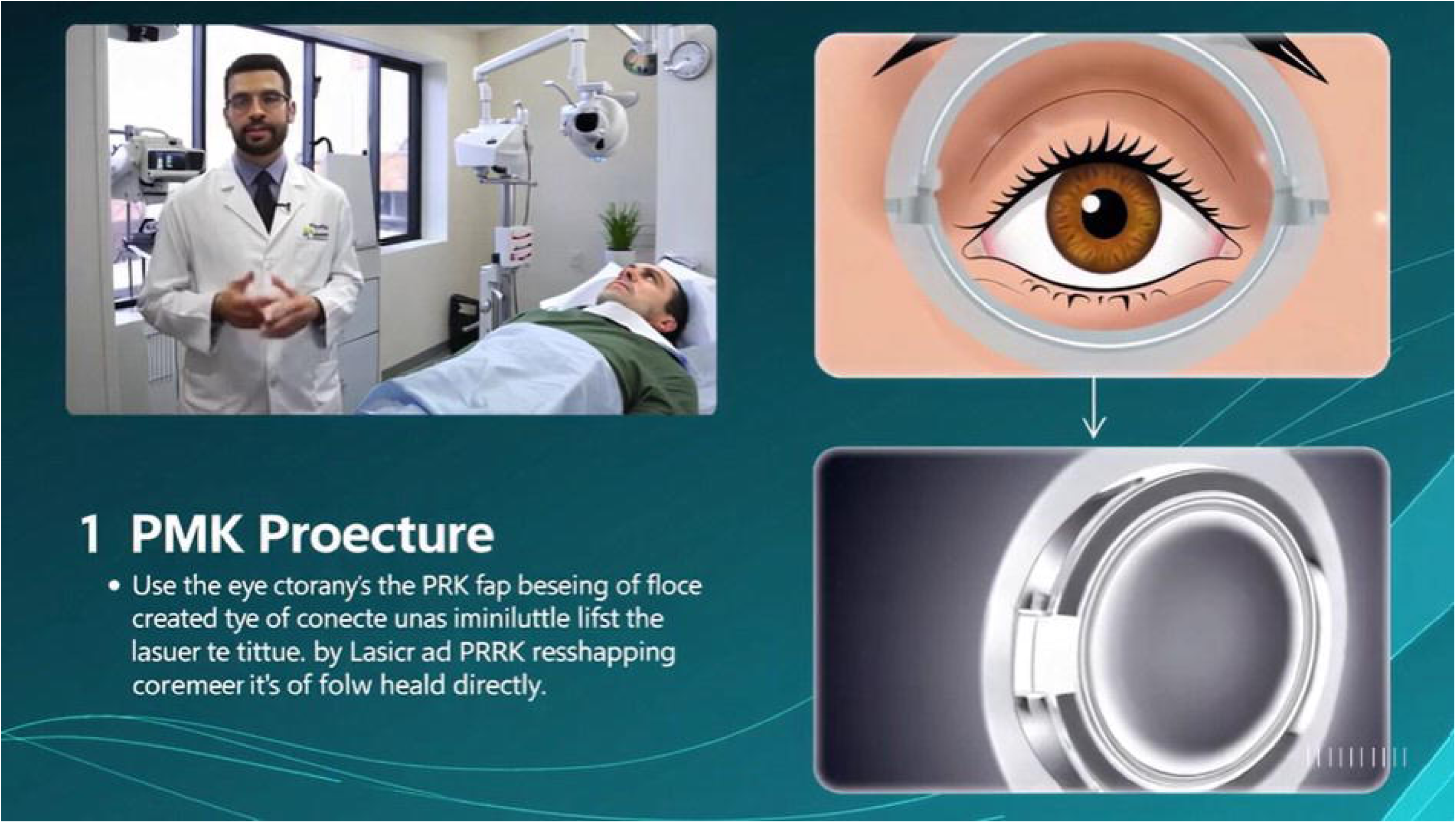

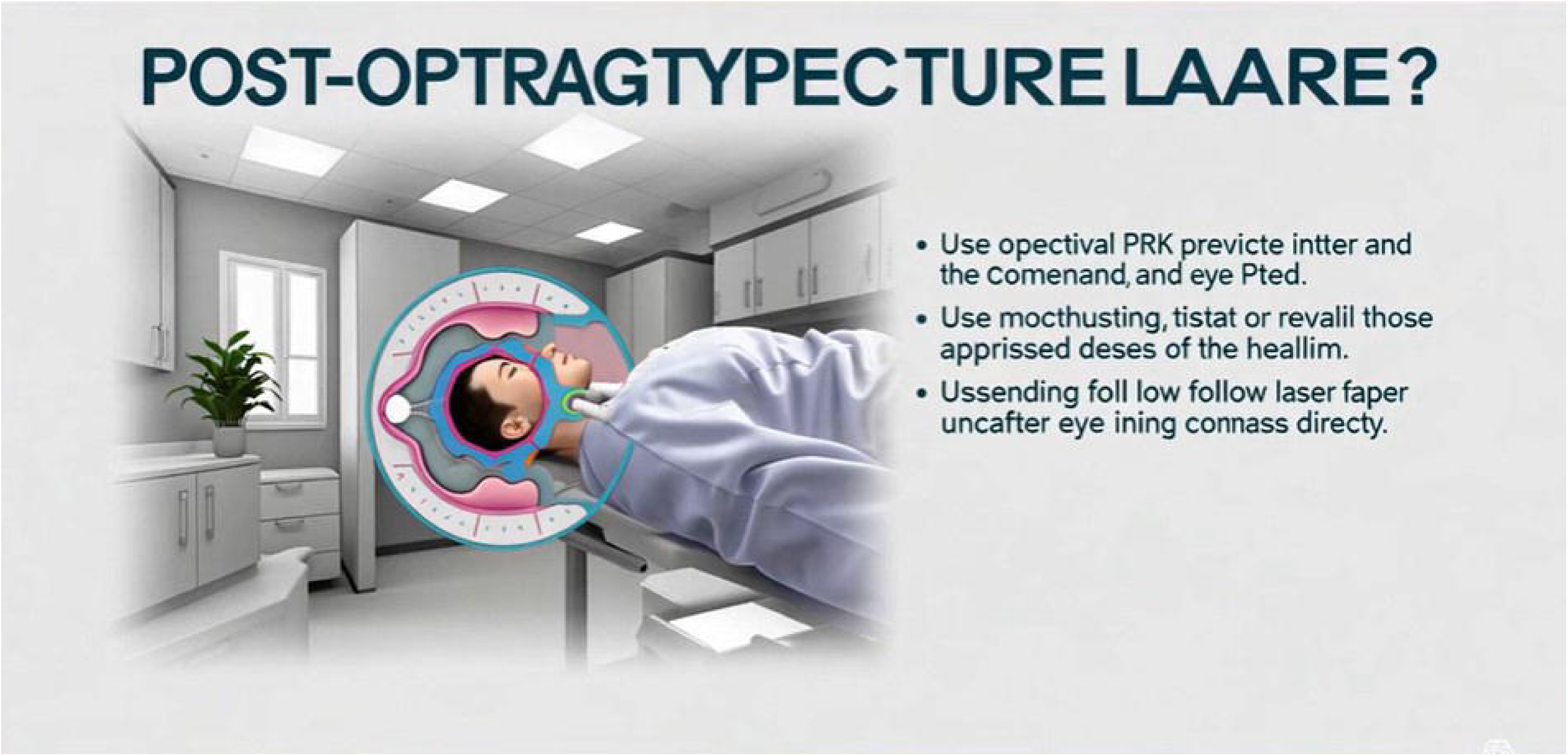

